# Level of Adherence to Recommended Lifestyle Modifications and Associated Factors Among Adult Hypertensive Patients Attending Chronic Follow-Up Units at Hypertension Sentinel Site Health Facilities - Bahir Dar City, Northwest Ethiopia, 2024

**DOI:** 10.1101/2025.01.14.25320524

**Authors:** Melaku Abebe, Kindie Fentahun Muchie, Tesfahun Taddege, Kebadnew Mulat

## Abstract

**Background:** Hypertension is one of the leading risk factors for mortality and morbidity worldwide. It is a chronic medical condition characterized by blood pressure (BP) consistently elevated above normal. Even though it is a global public health issue, it disproportionately affects populations in low- and middle-income countries that account for nearly 80% of deaths where health systems are weak. Adherence to medication therapy and lifestyle change is an aspect of patients’ care that is often overlooked and should be evaluated as a crucial part of cardiovascular management.

**Objective:** To assess the adherence to recommended lifestyle modifications and associated factors among adult hypertensive patients attending chronic follow-up units at hypertension sentinel site health facilities in Bahir Dar City, northwest Ethiopia in 2024.

**Methods:** A facility-based cross-sectional study was conducted in hypertension sentinel site health facilities in Bahir Dar city from April 24 to May 23, 2024. A total of 421 hypertensive patients were included in the study using a systematic random sampling technique. Data were collected through Computer-Assisted Personal Interviews (CAPI) and patient reviews using the KoboCollect. Data were analyzed using SPSS. Logistic regression analysis was used to identify factors associated with adherence to lifestyle modifications.

**Results:** The study found that 271 (64.4%) of the respondents were adherent to all studied lifestyle practices. Participants with good knowledge about lifestyle modification (AOR = 4.48 (2.30-8.75), good self-efficacy (AOR = 2.84 (1.61-5.01), good adherence to antihypertensive medication (AOR = 4.64 (2.54-8.45), primary education (AOR = 0.40 (0.18-0.89), underweight (AOR = 0.26(0.08-0.87), and overweight/ obese (AOR = 0.07 (0.04 -0.14) were factors associated with adherence to lifestyle modification.

**Conclusions:** The study revealed moderate level of adherence to lifestyle modifications among hypertensive patients. Good knowledge about lifestyle modification, good self-efficacy, and good adherence to antihypertensive medication were the factors positively associated with adherence to lifestyle modification, whereas, Patients with Primary education, underweight, and overweight/ obese were the factors negatively associated. Interventions targeting primary education, and underweight and overweight to raise knowledge status, medication adherence, and good self-efficacy will result in increased adherence.

## Introduction

Hypertension or raised blood pressure is one of the cardiovascular diseases(CVDs) characterized by blood pressure (BP) that is consistently elevated above normal(1). According to the Ethiopian national guideline on clinical and programmatic management of major NCDs, Hypertension is defined as SBP of 140 mmHg or higher, DBP of 90mmHg or higher, or both(2). In 90 – 95% of cases, the cause is unknown and it is called essential hypertension. Secondary hypertension refers to hypertension caused by other systemic illnesses as part of their manifestation(2). Hypertension is mainly classified as stage 1 hypertension (SBP 140-159 mmHg, or DBP 90-99 mmHg) and Stage 2 hypertension: SBP >160 mmHg, or DBP >100 mmHg. Hypertension is a leading risk for death and disability globally along with increasing co-morbidity(3).

Uncontrolled hypertension can cause irreversible harm to the kidneys, eyes, brain, and heart, and cause disability or even death(4). Hypertension management consists of pharmacologic interventions and lifestyle modification also known as non-pharmacologic management. Pharmacologic management entails the use of drugs from different classes to lower blood pressure(5). Unhealthy lifestyle choices characterized by unhealthy diets, heavy alcohol consumption, excess weight, and physical inactivity are the main risk factors for hypertension. Modification of these factors is associated with reductions in blood pressure(6). Lifestyle modification is thus an indispensable component in the management of hypertension; it lowers blood pressure, improves drug potency, and may even reduce or abolish the need for antihypertensive drugs(7).

The WHO recommended lifestyle modifications for hypertensive patients encompass weight reduction in those with a body mass index (BMI) greater than or equal to 25kg/m2, physical activity for at least 30 minutes on most days of the week, moderate alcohol intake, cessation of smoking and adoption of Dietary Approaches to Stop Hypertension (DASH) eating plan(8). The DASH eating plan is low in sodium, dairy fat, red meats and rich in fruit, vegetables, potassium, calcium, fish, and poultry(7). The protective effects of lifestyle modification span across all NCDs and delay the need for pharmacotherapy which is associated with adverse effects and is more costly.

Studies done in Ethiopia have identified risk factors related to hypertension, for example, being overweight, being of male gender(9); physical inactivity, cigarette smoking, adding salt to food in addition to the normal amount that is added to the food during cooking, history of diabetes(10); using animal product butter, and BMI 25.0 to 29.9 and greater than 30(11); alcohol drinking, and khat chewing(12); poor access and utilization of health services, absence of health insurance schemes, and rapid urbanization(13).

Hypertension, or high blood pressure, is a major public health concern worldwide, including in Ethiopia. If left uncontrolled, hypertension can lead to serious complications such as heart disease, stroke, and kidney failure. Hypertension is a major cause of premature death worldwide (14). It accounts for more loss of life from cardiovascular disease (CVD) than any other modifiable risk factor. More than 50% of people who lose their lives from coronary heart disease (CHD) and stroke have hypertension.

In Ethiopia, the prevalence of hypertension is increasing over time with a current prevalence of 21.8%, with higher rates among males (23.2%) compared to females (19.62%)(15). Uncontrolled hypertension is a risk for the occurrence of cardiovascular diseases such as stroke, myocardial infarction, and congestive heart failure as well as complications such as renal and eye diseases, stroke, and chronic kidney disease(16).

Poor adherence to lifestyle modifications can lead to suboptimal blood pressure control, increasing the risk of complications and diminishing the overall effectiveness of hypertension management strategies. Despite all its known adverse health consequences, high blood pressure is still poorly controlled globally due to low non-pharmacological interventions such as lifestyle modifications(16, 17). Despite the increasing burden of hypertension, there is very little data exists on the extent and factors related to adherence to the WHO-recommended lifestyle modifications, hence the need to investigate these.

There were few papers published in the country as well as in the study area on the topic. Some studies used the convenience sampling method which is prone to bias(18), some studies didn’t consider the independent factors such as knowledge about hypertension, social support and self-efficacy which are included in this study(18). Understanding the adherence levels to lifestyle modifications provides essential insights into how well patients are managing their hypertension beyond medication. This study can identify gaps in adherence that need to be addressed to enhance overall management and reduce the burden of the disease.

This study will also provide valuable information for health policy-makers in designing or redesigning treatment guidelines and protocols. It will also give insight for RTSL to know the impact of their program. Finally, this study will lay the groundwork for future research in the area of hypertension management, particularly in low- and middle-income settings. The insights gained could inspire further studies on intervention efficacy, long-term outcomes of adherence, and the integration of lifestyle modifications into routine clinical practice.

## Materials and Methods

### Study Area and Design

A facility-based cross-sectional study was conducted in hypertension sentinel site health facilities in Bahir Dar City, Amhara, Ethiopia. Bahir Dar is located in northwest Ethiopia from April 24 to May 23, 2024. It is the seat of the government of the Amhara National Regional State and is located 565 kilometres northwest of Addis Ababa, Ethiopia. The town’s total population (excluding rural Kebeles) is estimated to be about 332,856.19 The city administration is made up of 18 kebeles. There are seven hospitals (three public referral hospitals and four private hospitals). There are 62 sentinel primary health care facilities in 6 regions of the country, 7 of them found in the Amhara region. Shum Abo health center and Addis Alem primary hospital are the only two hypertension sentinel sites in Bahir Dar city.

### Populations

**Source population:** All hypertensive patients who attended the chronic follow-up unit

**Study unit:** Hypertensive patients who fulfilled the inclusion criteria and took part in the study.

### Eligibility Criteria

**Inclusion criteria:** All adult hypertensive patients on anti-hypertensive treatment for at least 6 months before the commencement of the study.

**Exclusion criteria:** Hypertensive patients who were critically ill and had mental health issues at the time of data collection were excluded.

### Sample size determination and procedure

The sample size was calculated using a single population proportion formula by considering the proportion of adherence to recommended lifestyle modifications at 46.4%(19), 95% CI, and 5% of marginal error. The sample size was estimated using the following assumptions. After adding a non-response rate of 10% to increase power, the total sample size was 421 participants. There are only two Hypertension sentinel sites in Bahir Dar city and both were taken conveniently for this study. The number of study units for each follow-up was proportionally allocated based on the number of patients coming per month and those who were part of the final sample size were selected using systematic random sampling.

### Data collection tools

The questionnaire included interview questions about socio-demographics, knowledge about the disease, and LSM adherence while clinical and treatment-related data were extracted from the patient chart and treatment follow-up registration book.

Patients’ adherence to LSMs was assessed using different tools from different resources including Hypertension Self-Care Activity Level Effects (H–SCALE), which has been validated for use in several studies(20). H-SCALE is a self-reported questionnaire that contains six categories of LSMs including medication adherence, low salt intake, smoking cessation, weight management, and alcohol abstinence. In this study, low salt intake, smoking cessation, and weight management were used. Questionnaires on Adherence to Alcohol moderation, Cessation of smoking, and physical exercise were prepared by the principal investigator according to JNC 8 recommendation. The categories of LSMs and other tools used are described as follows:

**Low-salt intake** contains ten items with each score of 0–7. Six questions were reversely coded. A mean score was calculated. Scores of 6 or better (indicating that participants followed low-salt diet practices on 6 out of 7 days) were considered adherent(17).

**Physical activity** was assessed by three-item questionnaires. The first question is about frequency of exercise in the week. The second is about the type of exercise and the final is on duration of the activities.

**Smoking habit** was assessed using two questions the first asks about the status of smoking and the second asks for the number of days of smoking if the respondent is a smoker(17).

**Weight management** was measured using a ten-item questionnaire based on a 5-point Likert scale ranging from 1 strongly disagree to 5 strongly agree, with a total score ranging from 10 to 50(17).

**Alcohol moderation: -** there were two questionnaires to assess alcohol moderation. A scoring scheme of Yes = 0 and No = 1 was used. One unit (drink) = a half pint of Tela/beer/lager (5 % alcohol), 100 ml of wine/Tej (10 % alcohol), Areke/spirits 25 ml (40% alcohol))(2).

**Adherence to anti-hypertensive medication**: - The 5-item Medication Adherence Report Scale (MARS-5) was used to assess patients’ medication adherence. The result was valued as always=0, often=1, sometimes=2, rarely=3, and never=4(21).

**Social support factors**:- Duke Social Support and Stress Scale with 12 items was used to assess support gained from family and non-family(22). Responses were valued as: “none” = 0, “some” = 1, “a lot” = 2, “yes” = 2, “no” = 0 and “there is no such person” = 0. Blank responses were considered as “0”.

**Knowledge about the disease:-** A modified 12-item knowledge assessment tool was used to assess participants knowledge about lifestyle modification(17). Responses were coded, “True” as “1” and “False” as “0” for questions One, Three, Six, Nine, Ten, and Eleven. Where reverse coding is used for the rest.

**Self-efficacy:-** Self-efficacy was assessed by the Chronic Disease Self-Efficacy Scale which contains 6 items(23). Responses were valued from One to Ten.

### Operational definition

**Adherence:** The extent to which a person’s behavior corresponds with recommendations from health care providers.

**Adherence to lifestyle modifications**: respondents who were adherent to all five (low salt diet, physical exercise, cessation of smoking, alcohol moderation, and weight management-related recommendations) recommended lifestyle modifications were considered adherent to LSM.

**Adherence to low-salt diet**: A mean score of 6 or better (indicating that participants followed low-salt diet practices on 6 out of 7 days were considered adherent.

**Adherence to physical exercise**: Participants who did moderate-intensity exercise for 30 min and above per day three times or above per week, or Participants who did vigorous-intensity exercise for 20 min and above per day three times or above per week.

**Adherent to smoking cessation** - Respondents who reported 0 days of smoking or 0 days of staying in a room/ ride in an enclosed vehicle while someone was smoking in the past seven days were considered nonsmokers.

**Adherent to weight management** - Participants who reported that they agreed or strongly agreed with all 10 items (score ≥40) were considered to be following good weight management practices.

**Adherent to alcohol moderation**: respondents who had reported that they either never consumed alcohol or who drank less than 2 units for women and 3 units for men were taken as adherent to alcohol moderation.

**Adherence to anti-hypertensive medications**: respondents who score 23 or more on the MARS-5 item scale of Medication Adherence are adherent to Medication.

**Co-morbidities**: respondents with one or more medical conditions in addition to hypertension.

**Knowledge about hypertension**: respondents with scores above the mean value on 12-item knowledge assessment tools were considered as having good knowledge about hypertension.

**Social support**: the support gained from family and non-family members. In this study, respondents whose score is above the mean value on the Duke Social Support and Stress scale were taken as having social support.

**Self-efficacy:** is the belief in one’s capabilities to organize and execute the courses of action required to produce a given attainment. In this study, respondents who scored above the mean value on the 6-item Chronic Disease Self-Efficacy Scale were considered to have good self-efficacy to cope with and manage their disease.

### Data management and analysis

Data were collected through Computer-Assisted Personal Interviews (CAPI) and patient reviews using the KoboCollect tool and the data was exported to Microsoft Excel 2021. Then the data was cleaned and exported to Statistical Package for Social Science (SPSS) version 27.0 for data analysis. Data was summarized using descriptive statistics (frequencies, percentages, means, and standard deviation). Binary logistic regression was used to identify factors associated with adherence to lifestyle modification among hypertensive patients in the study area. In the binary logistic regression, variables with a P-value ≤ 0.2 were entered into multivariable logistic regression and analysis by controlling confounding variables. The data was checked for multicollinearity and out-layer. Both Crude Odds Ratio (COR) and Adjusted Odds Ratio (AOR) with a 95 % confidence interval (CI) were used to show an association between adherence to lifestyle modification and selected variables. Variables having p-value ≤ 0.05 in the final model were assumed significant determinants. The model fitness test was evaluated by the Hosmer and Lemeshow goodness of fit test.

## Result

### Socio-demographic characteristics

During the study, 421 clients were planned to be included in the study and there was no refusal to participate in the study with a response rate of 100%. The study consisted of 219(52%) males. The mean age of the respondents was 55.2 ± 0.65 years while the majority of the respondents 221 (52.5%) were aged greater or equal to 56 years old. About 382(90.7%) of the respondents were Amhara by ethnicity. The majority of the respondents 331(78.6%) were orthodox by religion and 264(62.7%) were married. Out of the respondents, 342(73.4%) had formal education and 135 (32.1%) of respondents were government employees. Four hundred two (95.5%) respondents were urban residents and 187 (44.4%) of respondents had an income of ≤ 5000 Ethiopian Birr (ETB). Regarding social support for lifestyle modification, 221 (52.5%) of respondents had social support. The mean score for knowledge was 9.9 and out of the 421 participants, 327(77.7%) were found to be knowledgeable about hypertension. Regarding the average distance traveled to reach the hospital 299 (71%) of the respondents traveled less than 5KM.

**Table 1.** Socio-demographic characteristics of respondents attending chronic follow-up units at hypertension sentinel sites in Bahir Dar, Ethiopia, 2024.

| Variables | Categories | Frequency | Percentage (%) |
| --- | --- | --- | --- |
| Sex | Male | 219 | 52.0 |
|  | Female | 202 | 48.0 |
| Age | 18 – 35 | 20 | 4.8 |
|  | 36 – 45 | 124 | 29.5 |
|  | 46 – 55 | 56 | 13.3 |
| | $\geq 56$ | 221 | 52.5 |
| Religion | Orthodox | 331 | 78.6 |
|  | Muslim | 65 | 15.4 |
|  | Protestant | 25 | 5.9 |
| Marital status | Single | 42 | 10.0 |
|  | Married | 264 | 62.7 |
|  | Widowed | 65 | 15.4 |
|  | Divorced | 50 | 11.9 |
| Number of Children in the house | $< 3$ | 137 | 32.5 |
| | $> 3$ | 284 | 67.5 |
| Ethnicity | Amhara | 382 | 90.7 |
|  | Tigre | 20 | 4.8 |
|  | Oromo | 12 | 2.9 |
|  | Other | 7 | 1.7 |
| Educational level | Can't read and write | 23 | 5.5 |
|  | Read and write without formal education | 56 | 13.3 |
|  | Primary Education | 125 | 29.7 |
|  | Secondary Education | 128 | 30.4 |
|  | College/University | 89 | 21.1 |
| Occupation | Farmer | 65 | 15.4 |
|  | Governmental employee | 135 | 32.1 |
|  | Housewife | 68 | 16.2 |
|  | Merchant | 80 | 19.0 |
|  | non-governmental employee | 36 | 8.6 |
|  | Retired | 37 | 8.8 |
| Monthly income | < = 5000 ETB | 187 | 44.4 |
|  | 5001 - 9999 ETB | 127 | 30.2 |
|  | >= 10,000ETB | 107 | 25.4 |
| Residence | Rural | 19 | 4.5 |
|  | Urban | 402 | 95.5 |
| Social support for lifestyle practice | No | 200 | 47.5 |
|  | Yes | 221 | 52.5 |

### Health-related characteristics

Out of the total 421 respondents, 203 (48.2%) were 5 and above years since they were diagnosed with hypertension. One hundred thirty-five (32.1%) of the respondents had co-morbidities, of which, diabetes mellitus was found to be the most frequent comorbidity with 89 (67.9%) of the respondents having it; 31 (23%) had Asthma, (15) 11% had chronic kidney disease (CKD). Thirty (7.1%) of participants were underweight and 205(48.7%) were overweight/ obese. Of the participants, 327(77.7%) had good knowledge about LSM. Three hundred one (71.5%) participants have good adherence to medication.

**Table 2.** Health-related characteristics of respondents attending chronic follow-up unit of Hypertension sentinel sites in Bahir Dar City, Northwest Ethiopia, 2024.

| Variables | Categories | Frequency | Percentage (%) |
| --- | --- | --- | --- |
| Time since diagnosis of hypertension | < 1 Year | 26 | 6.2 |
|  | 1 - 5 Year | 192 | 45.6 |
|  | > 5 Year | 203 | 48.2 |
| Co-morbidities | Yes | 135 | 32.1 |
|  | No | 286 | 67.9 |
| Blood Pressure Status | Uncontrolled | 167 | 39.6 |
|  | Controlled | 254 | 60.3 |
| Number of Medication | < 2 | 270 | 64.1 |
|  | > 2 | 151 | 35.9 |
| Adherence to Medication | Good | 301 | 71.5 |
|  | Poor | 120 | 28.5 |
| Knowledge about HTN | Good | 327 | 77.7 |
|  | Poor | 94 | 22.3 |
| BMI | Normal | 186 | 47.0 |
|  | Underweight | 30 | 5.5 |
|  | Overweight/ Obese | 205 | 47.5 |
| Patient-provider relationship | Good | 24 | 5.7 |
|  | Not Good | 397 | 94.3 |
| Distance from HF | <= 5 KM | 299 | 71.0 |
|  | > 5 KM | 122 | 29.0 |
| Self-Efficacy | Good | 266 | 63.2 |
|  | Poor | 155 | 36.8 |
| Availability of Health Insurance | Yes | 73 | 17.3 |
|  | No | 348 | 82.7 |
| Availability of HTN drugs | Yes | 108 | 25.7 |
|  | No | 313 | 74.3 |

### Adherence to recommended lifestyle modifications

Of the 421 participants, 283 (67.2%) adhered to dietary changes, 292 (69.4%) followed weight management guidelines, and 309 (73.4%) engaged in regular physical exercise. Notably, 372 (88.4%) adhered to alcohol moderation recommendations. Smoking cessation is crucial for managing hypertension, and the study found that 421 (100%) adhered. **(Fig 1)**

**Fig 1.**
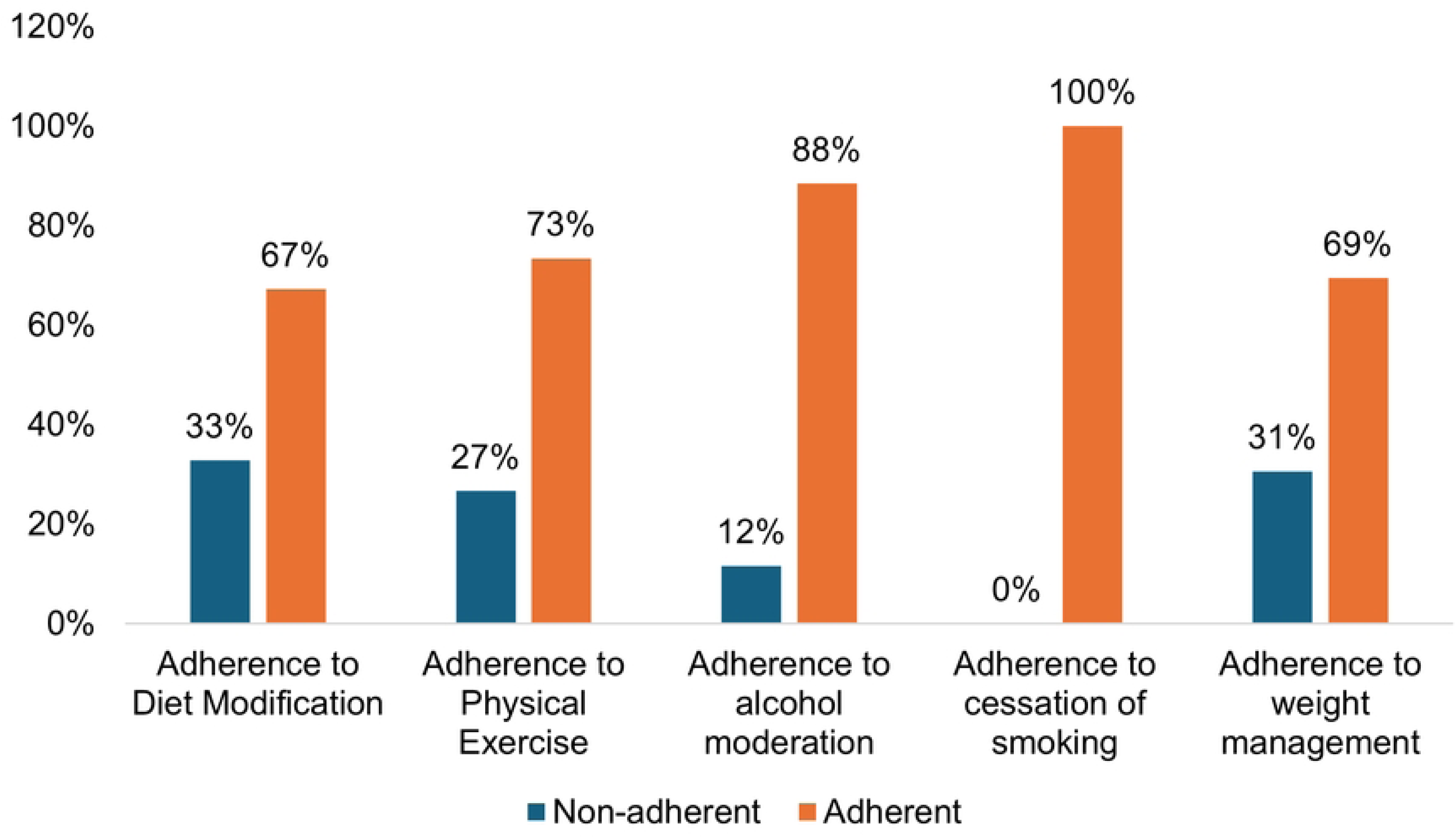
Adherence to Recommended lifestyle practice among hypertensive patients attending chronic follow-up unit of Hypertension sentinel sites in Bahir Dar City, Northwest Ethiopia, 2024

### Overall adherence to recommended lifestyle modifications

The overall adherence to recommended lifestyle modification practices, including exercise, diet, smoking cessation, weight management, and alcohol moderation, in the study was 64.4%. **(Fig 2)**

**Fig 2.**
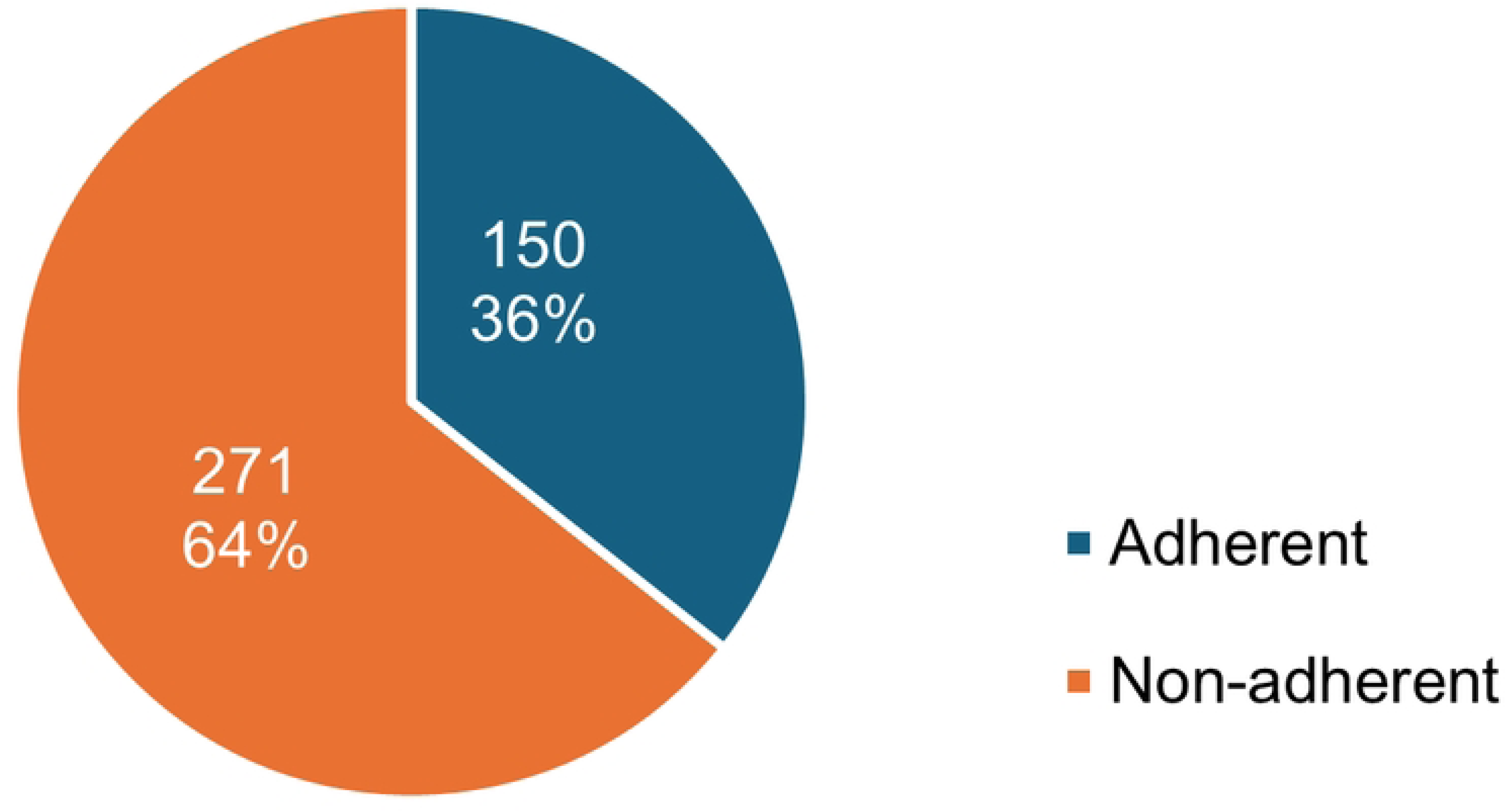
Overall adherence to lifestyle practice among hypertensive patients attending chronic follow-up unit of Hypertension sentinel sites in Bahir Dar City, Northwest Ethiopia, 2024

### Factors associated with adherence to lifestyle modification practice

Based on binary logistic regression analysis **Marital Status, Educational Status, Occupation, Monthly Income, Blood Pressure status, BMI, Comorbidity, Knowledge Status, Adherence to Medication, Level of Self Efficacy, Availability of Health Insurance, Distance from HF (12 variables)** were taken as candidates for multivariable logistic analysis.

**Table 3.**
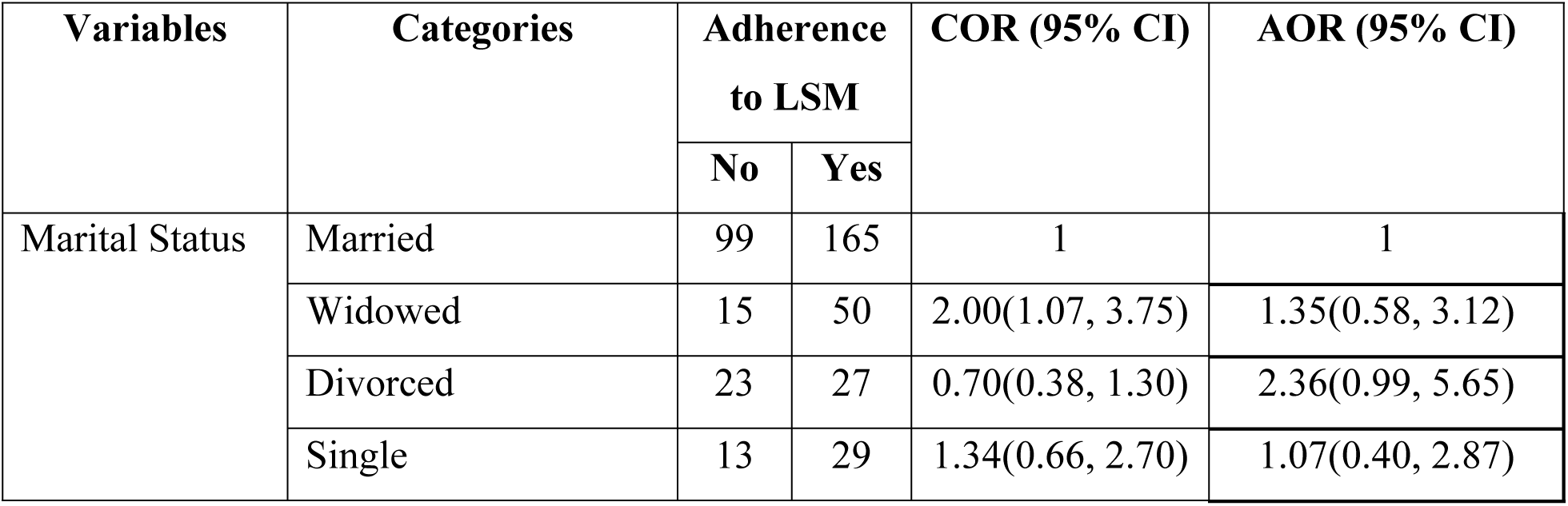
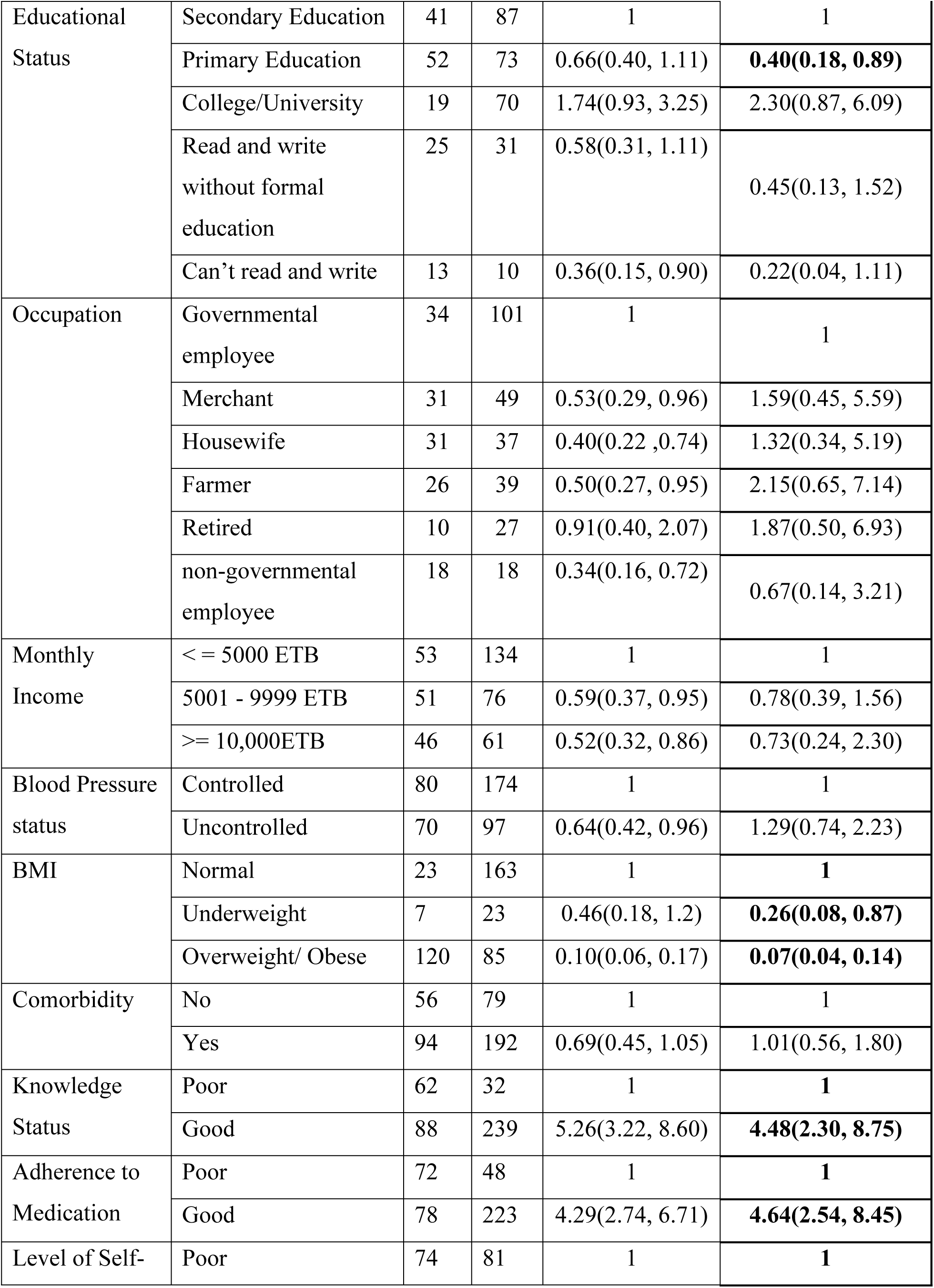
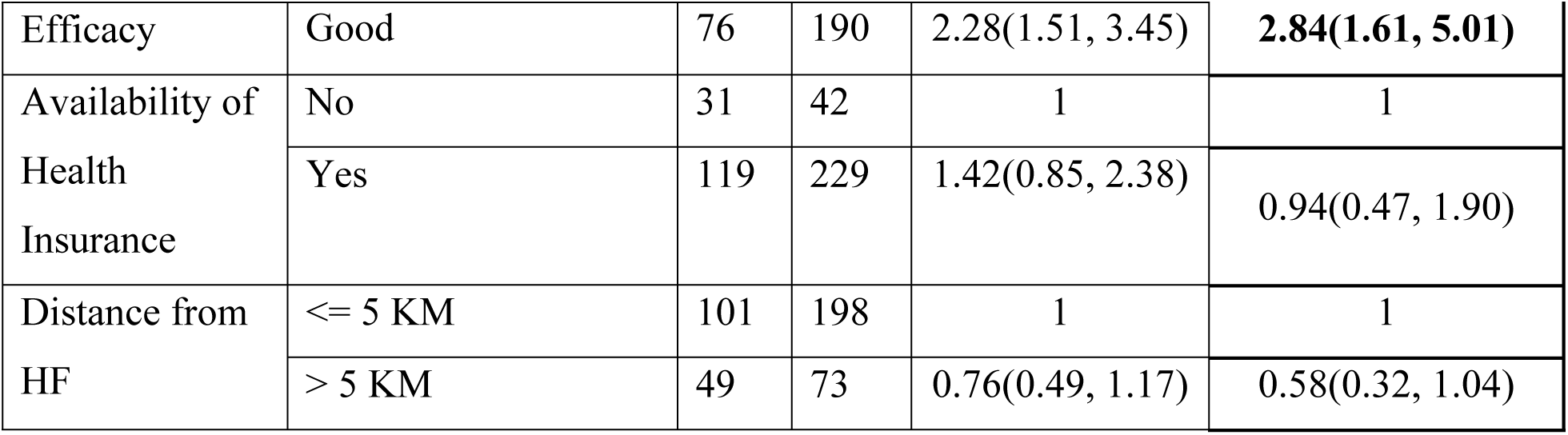
Bivariable and multivariable logistic regression showing actors associated with adherence to lifestyle practice among hypertensive patients attending chronic follow-up units of Hypertension sentinel sites in Bahir Dar City, Northwest Ethiopia, 2024.

According to the result of the multivariable analysis, educational status, BMI, knowledge status of the patient, medication adherence, and self-efficacy were independent variables that were associated with adherence to lifestyle modification practice among hypertensive patients. Educational status has a significant impact on the level of adherence to lifestyle modification (AOR = 0.40(0.18, 0.89)). BMI has a significant impact on adherence, underweight individuals (AOR = 0.26 (0.08, 0.87) and overweight/obese (AOR = 0.07 (0.04, 0.14) have a significant impact on adherence to LSM. Knowledge about hypertension significantly impacts adherence. Participants with good knowledge have a much higher likelihood of adherence (AOR = 4.48 (2.30, 8.75). Medication adherence strongly predicts adherence to lifestyle modifications. Those who adhere to their medication are significantly more likely to adhere to lifestyle modifications (AOR = 4.64 (2.54, 8.45). Self-efficacy is another critical factor. Participants with poor self-efficacy have significantly higher adherence rates (AOR = 2.84 (1.61, 5.01).

## Discussion

This study revealed that the magnitude of adherence to lifestyle modification in the selected health facilities was 271(64.4%). Where adherence to, low salt diet (67.2%), physical exercise (73.4%), alcohol moderation (88.4%), smoking cessation (1000%), and weight management (69.4%). According to the multivariable logistics regression, educational status, BMI, knowledge about lifestyle modification, self-efficacy, and medication adherence were strongly associated with adherence to lifestyle modification.

The overall adherence to LSM is higher than the findings reported from Nigeria (26.8%), Arbaminch (40.7), Addis Ababa Yekatite 12 Hospital (45.4%), Oromo Special Zone (52.7%), and another study done in Bahir Dar city (32.4%)(24–28). Our finding is lower than the findings of the study done in Nepal (79.2%) and Ghana (72%)(25, 29). This inconsistency might be sample size differences and differences in population characteristics. Another reason could be different exposure to lifestyle information and tools used to measure the lifestyle practice.

In this study, diet-related adherence was 67.2%. This finding is in line with another study done by Adiss Ababa public health hospitals (69.1%)(30). This finding is higher than studies reported from Nepal (30.3%), Bahir Dar City (48%), and Mizan Tepi University Teaching Hospital (60.8%)(28, 29, 31). The finding is lower than studies reported from Nigeria (94.4%), Oromo Special Zone (80.3%), Arbaminch (73.7%), and Dessie Referral Hospital (76.4%)(19, 24, 26, 32). This discrepancy may be due to differences in dietary habits and knowledge about the effect of a salt diet on blood pressure management.

This study revealed that 73.4% of participants adhered to regular physical exercise, which was held 3 or more days per week and 30 minutes or more duration. This result is less than studies reported from studies reported from Nepal (57.9%), public health hospitals in Addis Ababa (31.4%), Bishoftu town (29.8%), Arbaminch (29.3%), Dessie (39.9%), Mizan Tepi University Teaching Hospital (62.6%)(19, 26, 29, 30, 33).

In our study adherence to alcohol moderation is 88.4%. This finding is in line with studies done in Nigeria (90.73%), Bishoftu (87.9%), and Arbaminch (84.1%)(25, 26, 33). This finding is higher than studies done in Bahir Dar city (63.3%), Oromo Special Zone (51%), Mizan Tepi University Teaching Hospital (43.9%), public health hospitals in Addis Ababa (74.8%)(24, 28, 31, 32). This finding is less than studies reported from Nepal (95.1%), Nigeria (97.4%), and Dessie Referral Hospital (94%)(19, 25, 29).

In this study, all participants adhered to smoking cessation. This result is in line with studies done in Nigeria (100%)(34). There was a lower percentage of adherence reported compared to our result from Nepal 92.8%, Bishoftu (49.2%), Mizan Tepi (62.6%), public health hospitals in Addis Ababa (85.9%), Dessie Referral Hospital (89.7%)(19, 29, 31, 33). This high adherence rate to smoking cessation could be due to sociocultural norms that discourage smoking.

In our study, 69.4% of participants adhered to weight management. This study is comparable with a study done in Selected Hospitals in Central Gondar Zone (65%) and selected hospitals in Eritrea (68.9)(35, 36). The result is higher than studies reported from Dessie referral hospital (46.7%), Bahir Dar city (48.7%), Jimma University Specialized Hospital (43.1%)(19, 28, 37).

According to this study educational level(p=0.021) has a statistically significant association with adherence to LSM. This finding is in line with a study done in Nepal(p=0.021), Saudi Arabia(p=0.002), Africa(p=0.01), Bishoftu (p=0.037) and Mizan Tepi university teaching hospital (p ≤ 0.05)(29, 31, 33, 38, 39). Whereas studies done in Jordan (p = .220), Nigeria (p = .213), and the Oromo special zone(p>0.05), Felege Hiwot Referal Hospital (Bahir Dar) (0.089) found no association b/n the two variables(24, 32, 40).

According to this study, BMI emerges as a significant predictor of adherence to lifestyle modifications among patients with hypertension. Underweight individuals are 74% less likely to adhere to lifestyle compared with normal participants with BMI, whereas individuals who are overweight or obese are 90% less likely to adhere to lifestyle modification compared to normal BMI. This result is in line with studies done in Japan by Matsushita et al. (2018)(41). Whereas studies done in Jordan showed no association b/n body mass index and adherence to lifestyle modification(40, 42).

In this study participants with good knowledge about lifestyle modification are found to be 4.48 times more likely to exhibit adherence compared to those with poor knowledge. This finding was supported by studies from Jordan (AOR =2.96, CI = 1.037-8.45), Bahir Dar city hospitals (AOR =3.323, CI = 1.79-6.17), and Dessie Referral Hospital (19, 28, 40). This is because knowledge of hypertension lifestyle modification practices helps have a positive impact on individual patients’ access, utilization, and outcomes of the recommended lifestyle practices. A higher number of predictions was reported from public health hospitals in Addis Ababa (AOR =13.26, CI = 4.12-42.71), Bishoftu Town Public Health Facilities (AOR =8.22, CI = 3.87-17.47)(30, 33). The discrepancy may be due to different study populations or better information and living standards in the case of Addis Ababa.

Participants who had good self-efficacy were 2.84 likely to be adherent to lifestyle modification among the participants. This result is comparable with studies done in public hospitals in Addis Ababa (AOR =3.92, CI = 1.61 – 9.56), Bahir Dar city hospitals (AOR =3.553, CI = 1.910 – 6.513), Dessie Referral Hospital (AOR =3.64, CI = 1.75 – 7.55), Jimma University Specialized Hospital (AOR = 3.74, CI = 2.21-6.32) (28, 30). The possible explanation might be that the more confident patients are the more motivated and have emotional well-being to modify their lifestyle based on the recommendations, the more likely they are to be adherent.

Participants who adhere to their medication regimen are 3.74 times more likely to exhibit adherence to lifestyle modifications compared to those who adhere to their medication (AOR = 3.74, CI = 2.21-6.32). The result is in line with the finding from Yekatit 12 Hospital Medical College (AOR = 2.13, CI = 1.38-3.28)(27).

## Conclusion and recommendations

This research aimed to assess the level of adherence to lifestyle modification and associated factors among hypertensive patients in hypertension sentinel sites in Bahir Dar City, Ethiopia. The study revealed a moderate level (64.4%) of adherence to lifestyle modifications among hypertensive patients. Healthcare providers should focus on educating patients about the importance of lifestyle modifications, particularly for those with risk factors such as abnormal BMI, poor knowledge, and poor medication adherence. Interventions targeting these factors may improve adherence and

## Strengths and limitations of the study

During the study time, health education was given by for each of the study participants about the benefits of practicing lifestyle modification for controlling blood pressure. The data was self-reported by the participants; there may be a social desirability bias and recall bias, which affects the result of the study. It was difficult to assess the amount of salt intake of the patients. Since the study is done in hypertension sentinel sites the result doesn’t represent non-sentinel sites.

## Data Availability

The data underlying the results presented in the study are available from the corresponding author, Melaku A., upon reasonable request. He can be contacted via email at.

## Abbreviations

ACC: American College of Cardiology
AHA: American Heart Association
AOR: Adjusted Odd Ratio
BMI: Body Mass Index
BP: Blood Pressure
COR: Crude Odd Ratio
CVD: Cardio Vascular Disease
DASH: Dietary Approach to Stop Hypertension
HTN: Hypertension
HELM: Hypertension Evaluation of Life Style Modification
PIH: Pregnancy Induced Hypertension
WHO: World Health Organization.

## Author contributions

Conceived and designed the study, performed analysis and interpretation of data, drafted the manuscript, and finalization. KM, KF, and TT assisted with the design conception, analysis, and interpretation of data and the critical review of the manuscript. All authors read and approved the final manuscript.

## Authors’ details

Two of the authors are academicians at Bahir Dar University and the other one works at USAID/Ethiopia Health Resilience Activity, MSH-Ethiopia. MA has an MPH in Field Epidemiology, KM has an MPH in Epidemiology, KF has a PhD in Biostatistics, and TT has an MPH in Field Epidemiology and RH.

## Acknowledgment

We would like to acknowledge Bahir Dar University College of Medicine and Health Sciences, School of Public Health, hospital admins, data collectors, and patients who participated in the study.

## Availability of data and material

The data is available on the corresponding author Melaku Abebe Melaku upon request.

## Consent for publication

Not applicable

## Ethics approval and consent to participate

The Bahir Dar University College of Medicine and Health Sciences Institutional Review Board (IRB) approved the protocol on the ethical principles of the Helsinki Declaration and assigned it reference number 886/2023 **(S1 Fig.)**. APHI also provided a letter granting permission for the facilities to take part in the study, with letter number 03/2106. For the patients’ willingness to participate, written informed consent was obtained from them. All the procedures that included human participants adhered to the Declaration of Helsinki.

## Supporting Information

S1 Fig. Bahir Dar University College of Medicine and Health Sciences Institutional Review Board (IRB) Decision

